# Gambling Disorder: Simultaneous Measurement of Screening Scales in Multiple Regions of Japan

**DOI:** 10.1101/2022.11.27.22282778

**Authors:** Tatsuya Noda, Moritoshi Kido, Chieko Ito

## Abstract

**Background and Objectives:** Gambling disorder is defined as persistent and recurrent problematic gambling behavior leading to clinically significant impairment or distress in DSM-5 diagnostic criteria. According to the DSM-5, the past-year prevalence rate of gambling disorder is about 0.2%-0.3% in the general population, and the lifetime prevalence is about 0.4%-1.0%. Japan currently has no casinos, but three integrated resorts (IRs), including casinos, are scheduled to open in the late 2020s. In abroad, there have been studies on the effect of casinos on the prevalence of gambling in a region, but similar empirical studies do not exist in Japan. There are a lot of surveys being conducted in many areas, but the different screening tests used make it difficult to compare the results. However, comparisons between screening tests are less common. The first objective of this study is to measure the prevalence of gambling disorder in areas where IRs are planned to open and control areas. The second objective is to identify relationships among screening tests and examine differences between measures by administering multiple screening tests for gambling disorder to the same subjects.

**Materials and methods:** This survey was a self-administered, online survey of 2000 people in Japan, ranging in age from 20 to 69 conducted in 2021. We administered 4 gambling disorder screening tests (SOGS, PGSI, LieBet Screen,DSM-5). Experience rates for each of 8 gambles were measured. Percentage above cut-off score (PAC) was calculated and compared by gender and region for 4 tests.

**Results:** Among the eight gambling activities, the lottery had the largest experience rate at 58.5%, PACHINKO was second at 42.2%, and there were no major regional disparities. In each of the 4 screening tests, PAC was greater in males, and regional differences were found in SOGS and DSM-5. The distribution of PAC for different cut-off scores for each test is illustrated in a “dango chart” for comparison.

**Discussion:** This study indicates PACs for 4 gambling disorder screening tests in planned and non-IR areas, and this will be a baseline survey to measure the impact of opening an IR. Although our results will contain some biases likely due to methodological factors, by continuing to use the same measurement method, our research project will be able to reach its ultimate goal of observing changes in the applicable rate before and after the opening of the IR. In addition, this study administered several gambling disorder screening tests to the same subjects in the same region and illustrated them in a chart format named the dango chart. This visualized the changes in the applicable person rate when the cut-off scores of the screening tests were changed (within-test comparisons) and the differences in the distribution of the applicable person rate across tests (between-test comparisons). We think that the dango chart is a useful presentation method for comparing several different tests.

## 1. Introduction

### 1.1. Gambling disorder

A gambling disorder first appeared in the third edition of the Diagnostic and Statistical Manual of Mental Disorders (DSM-III) published by the American Psychiatric Association (APA) in 1980 using the diagnostic name “pathological gambling” (Petry & Blanco, 2013). Currently, the DSM-5 has diagnostic criteria of gambling disorders in the non-substance-related disorders group of substance-related and addictive disorders. The DSM-5 diagnostic criteria define gambling disorder as “persistent and recurrent problematic gambling behavior leading to clinically significant impairment or distress” and that “the gambling behavior is not better explained by a manic episode”. The DSM-5 has nine diagnostic criteria for gambling behavior, and a person is diagnosed as gambling disorder if four or more of the criteria are met for gambling behavior in the past 12 months.

### 1.2. Prevalence of gambling disorder

The prevalence of gambling disorders varies widely by region and survey methods. According to the DSM-5, the past-year prevalence rate of gambling disorder is about 0.2%-0.3% in the general population, and the lifetime prevalence is about 0.4%-1.0% (APA, 2013). Williams, Volberg & Stevens (2012) collected studies of prevalence surveys from 1975 to 2012 and compared the results. And they compared the period prevalence rates from each study by weighting and modifying the prevalence rates based on differences in the scales used, the time frame, what the subjects were classified as above the cut-off scores, the survey method, and when the scales were administered.

The results indicate that the past-year prevalence rate is 0.5%-7.6%. The results also show regional characteristics, with Europe having the lowest, North America and Australia having moderate, and Asia having the highest. Calado & Griffiths (2016) reviewed studies of prevalence surveys conducted in a total of 30 countries from 2000 to 2015, collecting a total of 69 studies from North America (3 studies), South America (1 study), Asia (10 studies), Oceania (5 studies), Africa (4 studies), and Europe (46 studies). The results of these studies indicate that the past year prevalence rate ranges from 0.1%-5.8% and lifetime prevalence rate from 0.7%-6.5%. By region, the past-year prevalence rate was shown to be 2.0%-5.0% in North America, 0.5%-5.8% in Asia, 0.4%-0.7% in Oceania, and 0.1%-3.4% in Europe. These previous studies have shown that the prevalence rate in Asia is higher and more varied than in other regions. However, this is limited to data from regions with casinos, and the Asian prevalence does not include Japanese results.

An epidemiological study about the prevalence rate of gambling disorder in Japan was conducted by the Agency for Medical Development of Japan (AMED) in 2017. From the results of the South Oaks Gambling Screen (SOGS) (Lesieur & Blume, 1987), a screening test for gambling disorder, it was indicated that the past-year prevalence rate in Japan was 0.8% and the lifetime prevalence rate was 3.6% (Higuchi & Matsushita, 2017). Recently, a national prevalence study was conducted in 2020 and results from the SOGS and Problem Gambling Severity Index (PGSI) revealed a point prevalence of 2.2% (SOGS) and 1.6% (PGSI), respectively(Matsushita, Nitta, & Toyama, 2021). Previous studies collectively indicate a range of 0.1%-7.6% with the past-year prevalence rate (Williams, Volberg & Stevens, 2012 ; Calado & Griffiths, 2016), while Asia has a higher prevalence rate compared to other regions. In addition, the results of a fact-finding survey indicate a point prevalence of 0.8% and a lifetime prevalence of 3.6% in Japan.

### 1.3. Screening tests used in prevalence studies

According to Williams, Volberg & Stevens (2012), the screening tests used in the 1975-2012 prevalence study were as follows:

- SOGS for 103 studies,
- Diagnostic and Statistical Manual of Mental Disorders (DSM) (DSM-III published in 1980; DSM-III-Revised in 1987; DSM-IV in 1994) for 64 studies.
- Canadian Problem Gambling Index (CPGI; Ferris &Wynne, 2001) for 55 studies.
- Other screening tests for 20 studies.

In addition, according to Calado & Griffiths (2016), the screening tests used in prevalence studies conducted between 2000 and 2015 were as follows:

- SOGS for 23 studies.
- Problem Gambling Severity Index (PGSI; Ferris &Wynne, 2001) for 21 studies. PGSI is consisted of 9 items from CPGI.
- DSM-IV for 21 studies.
- the National Opinion Research Center DSM Screen for Gambling Problems (NODS) (Gerstein et al., 1999) for 7 studies.
- the Diagnostic Interview Schedule for pathological gambling (DIS) (Robins, Marcus, Reich, Cunningham, & Gallagher, 1996) for 3 studies.
- Gamblers Anonymous Twenty Questions (GA20) (Gamblers Anonymous, 1984) for 3 studies.
- the Lie/Bet scale (Johnson et al., 1997) for 2 studies.
- the Diagnostic Interview for Gambling Severity (DIGS) (Stinchfield, 2002) for 1 study.

These two results indicate that SOGS is the most commonly used screening test and CPGI/PGSI is the second most commonly used screening test. Williams, Volberg & Stevens (2012) also show a trend that SOGS was used for most prevalence studies until 2000, but since 2001, CPGI and DSM have been replacing SOGS in use.

### 1.4. Problems with Prevalence Surveys and Factors Making Comparisons Between Prevalence Surveys Difficult

As explained in the previous section, prevalence studies of gambling disorder have been conducted in various countries and regions, and a lot of data including period prevalence rate and lifetime prevalence rate was obtained. However, these prevalence studies have the following methodological differences:

- Screening tests employed
- Cut-off scores setting
- The time frame of the prevalence at the point in time (e.g., past 12 months or 6 months)
- Prevalence being measured (lifetime, point in time)
- Differences in the method of administration (in-person interview, telephone interview, mail survey, online survey research).

These differences make it difficult to simply compare prevalence rates across studies (Williams, Volberg & Stevens, 2012). Regarding this issue, Williams, Volberg & Stevens (2012) used methodological differences among the studies, particularly differences in the screening tests used, lifetime or point in time prevalence, survey methods such as telephone or mail surveys, and description of the survey to the survey participants, to weights and results of previous prevalence surveys were compared by calculating standardized prevalence rates. A national prevalence study conducted in Japan in 2017 showed a period prevalence of 0.8% (1.5% for men and 0.1% for women) and a lifetime prevalence of 3.6% (6.7% for men and 0.6% for women) from face-to-face interviews using SOGS (Higuchi and Matsushita, 2017). The 2020 national prevalence study was conducted by mail and online, with SOGS results indicating a period prevalence of 2.2% (3.7% for men and 0.7% for women) and PGSI results indicating a period prevalence of 1.6% (2.8% for men and 0.4% for women) (Matsushita, Nitta, & Toyama, 2021). Local governments in Japan have also conducted their own surveys about the prevalence of gambling disorders. In 2018, Chiba City conducted an online survey using PGSI and found a period prevalence rate of 7.2% for men and 1.2% for women (Chiba Center for Mental Health, 2018). In 2020, a mail survey using SOGS was also conducted in Kanagawa Prefecture, obtaining data for a period prevalence rate of 0.8% and a lifetime prevalence rate of 4.9% (Kanagawa Prefecture, 2020). In the same year, Yokohama City conducted an interview survey using SOGS and obtained data for a period prevalence rate of 0.5% and a lifetime prevalence rate of 2.2% (Yokohama City, 2020).

However, each of these surveys has different survey procedures: the 2017 national prevalence study and the Yokohama City survey used interviews using SOGS, the Chiba City survey used online surveys using PGSI, the Kanagawa Prefecture survey used mail surveys using SOGS, and the 2020 national prevalence study used mail and online surveys using SOGS and PGSI. A simple prevalence comparison is not possible, as Williams, Volberg & Stevens (2012) pointed out that different screening tests used and different survey methods conducted can result in different prevalence rates in the same target population. In addition, PGSI has been administered more frequently than SOGS in recent years, which raises the question of consistency with past results when the screening test used is switched.

Therefore, in order to compare various prevalence studies, we need to conduct a survey using various screening tests and compare the results in Japan as in the study by Williams, Volberg & Stevens (2012).

### 1.5. Social Background of Gambling Disorder Research in Japan

Japan currently has no casinos, but three integrated resorts (IRs), including casinos, are scheduled to open in the late 2020s. And there is an active debate going on as to whether the introduction of IRs will increase the prevalence of gambling disorders.

While there have been studies on the effect of casinos on the prevalence of gambling in a region (Williams, West & Simpson, 2012), similar empirical studies do not exist in Japan. It is necessary to measure the regional prevalence before the opening of casinos in Japan for the purpose of conducting empirical research on the impact of casino opening.

### 1.6. Objective

The first objective of this study is to measure the prevalence of gambling disorder in areas where IRs are planned to open and control areas. The second objective is to identify relationships among screening tests and examine differences between measures by administering multiple screening tests for gambling disorder to the same subjects.

## 2. Materials and methods

### 2.1. Survey subjects

In this study, Osaka Prefecture (population 8.8 million), where IR is planned, and Fukuoka Prefecture (population 5.12 million), an urban area similar to Osaka Prefecture but with no plans to attract IR at the time of the survey, were selected as control areas where the impact of gambling measures implemented by areas planning to attract IR would be small. At the time of the survey, Wakayama Prefecture, Sasebo City in Nagasaki Prefecture, Yokohama City in Kanagawa Prefecture, Tokyo Prefecture, and Nagoya City in Aichi Prefecture were planning or considering the attraction of IR, all of which are a distance from Fukuoka Prefecture. The survey was a self-administered, online survey of 1,000 people each from Osaka and Fukuoka prefectures, ranging in age from 20 to 69, who were registered on the survey panel of Asmark Co. Due to the age structure of the survey panel, the maximum age for the survey was set at 69.

### 2.2. Survey instruments

A total of 57 items were used in the survey. It consists of the following questions:

- The items included basic attributes such as gender, age, place of residence, marital status, and number of people living together (6 items)
- Gambling experience (8 items)
- Maximum amount of money spent on gambling (per day/month) (2 items)
- SOGS (13 items)
- PGSI (9 items)
- LieBet Screen (2 items)
- DSM-5 (9 items)
- The items related to subjective economic status (1 item)
- The items related to the relationship between gambling and suicide attempts (1 item)
- K6 (6 items).

#### 2.2.1. Items about Basic Attributes

Respondents were asked about their gender (2 options), age (ratio scale), prefecture (47 options), municipality (open-ended), marital status (4 options), and number of people living together (ratio scale).

#### 2.2.2. Items about gambling experience and the maximum amount of money spent on gambling

Asked about their experience with legal gambling that currently exists in Japan (horse race, bicycle race, boat race, motorcycle race, lottery, sports promotion lottery, PACHINKO and casino outside Japan) using the 5-point Likert scale: “1-Never”; “2-Less than once a week within a year”; “3-More than once a week within a year”; “4-Less than once a week more than a year ago”; “5-More than once a week more than a year ago”. The question about the maximum amount spent on gambling asked about the maximum amount spent in a day and the maximum amount spent in a month on gambling during the year using 9 options; “1-I have not gambled at all in the past year”; “2-less than 100 yen”; “3-101 to 1,000 yen”; “4-1001 to 5,000 yen”; “5-5001 to 10,000 yen”; “6-10001 to 100,000 yen”; “7-100,001 to 1 million yen”; “8-1 million 1 or more yen”; “9-I do not know”.

#### 2.2.3. SOGS

There were a total of 12 screening items related to SOGS scores, and these items were about description of past gambling behavior :

- Yes / No questions (8 items)
- Choose one of four options: “1. never”, “2. some of the time (less than half the time) I lost”, “3. most of the time I lost”, “4. every time I lost” (1 item)
- Choose one of Three options:”1. never (or never gamble)”, “2. yes, less than half the time I lost”, “3. yes, most of the time”(1 item), “1. no”, “2. yes, in the past, but not now”, “3. yes”(1 item)
- Multiple-choice item: “a. from household money”, “b. from your spouse”, “c. from other relatives or in-laws’’, “d. from banks, loan companies, or credit unions’’, “e. credit cards’’, “f. loan sharks”, “g. you cashed in stocks, bonds, or other securities”, “h. you sold personal or family property”, “i. you borrowed on your checking account”, “j. never borrowed money to gamble”(1 item)

Scores ranged from 0 to 20, with a total score of 5 or more points indicating a probable pathological gambler (Lesieur & Blume, 1987). When translating the SOGS screening items, the author used the back-translation method, in which the English is translated into Japanese, the Japanese is translated back into English, and the Japanese translation is compared with the original English and examined.

#### 2.2.4. PGSI

The PGSI consisted of 9 items about one’s gambling behavior in the past 12 months. Responses were scored on a four-point Likert scale: “1. never”, “2. sometimes”, “3. most of the time”, “4. almost always”. Scores ranged from 0 to 27, the total score is classified as follows: 0 = non-problem, 1-2 = low risk, 3-7 = moderate risk, and 8 or more = problem gambling (Ferris & Wynne, 2001). In this study, we used the Japanese version of the PGSI (So, Matsushita, Kishimoto & Furukawa, 2019).

#### 2.2.5. Lie Bet Screen

The Lie Bet Screen consisted of two items about one’s gambling behavior in the past year. Responses were scored on Yes / No questions. Scores ranged from 0 to 2, with 0 indicating a non-problem and 1 or more indicating further assessment is needed (Johnson, Hamer & Nora, 1997). This study used the Lie Bet Screen items translated into Japanese by the JGSS (JGSS Research Center, Osaka University of Commerce, 2017).

#### 2.2.6. DSM-5

The DSM-5 diagnostic criteria for gambling disorder consisted of 9 criteria about one’s gambling behavior in the past 12 months. Those who meet four to five criteria are considered mild, six to seven are considered moderate, and eight to nine are considered severe (APA, 2013). In this study, we used the Japanese version of the diagnostic criteria of the DSM-5 (APA, 2017; Takahashi & Ohno, 2014, p.578). Responses were scored on Yes / No questions.

#### 2.2.7. Other Items

Other items included 1 item about subjective economic status, 1 item about oneʼs gambling behavior and suicide attempts, and items asking about psychological problems related to depression in the past 30 days (K6: 6 item). An item about subjective economic status was answered on a five-point scale: “1. very difficult,” “2. somewhat difficult,” “3. normal,” “4. somewhat comfortable,” and “5. very comfortable”. An item about suicide attempts was answered Yes / No. The K6 was scored on a five-point Likert scale: “1. not at all”, “2. a little”, “3. sometimes”, “4. usually”, “5. always”. Scores ranged from 0 to 24, with 13 or more points indicating severe mental disorder (Furukawa, et al., 2008).

### 2.3. Survey procedure

This online survey was conducted in April 2021 among those registered as survey monitors with ASMARQ Co. In this survey, those who were registered on the survey monitor website and resided in Osaka and Fukuoka prefectures were targeted and sent an e-mail with a survey request. The email included a description of the survey, its purpose, and a URL for the survey. After confirming the survey content and agreeing to the purpose, the subjects clicked on the URL link and responded to the survey.

Smartphones, personal computers, and tablets were used as tools to answer the survey. The number of survey targets was set at 1,000 for each prefecture, and the survey for a given sex and age group was considered complete when it reached the assigned sex and age group population, which was set to be the same proportion as the sex and age group population of each prefecture. In addition, subjects who completed the survey were paid a reward by Asmark for their responses.

### 2.4. Terminology

In this study, the number of persons who fit each score on the screening scale for gambling disorder was expressed as the number of applicable persons, and the percentage of applicable persons as a proportion of the total was expressed as the percentage of applicable persons. The Percentage Above Cut-off score (PAC) is the cumulative percentage of persons who fit the cut-off scores for a given score or higher. PAC represents the “prevalence,” so to speak, of adopting any score as the cut-off scores. However, since this study used a screening scale, it is not possible to say with certainty whether a client with a score above a certain level is truly a patient with a gambling disorder. In order to maintain neutrality in the interpretation of the results, the term “prevalence” is not used in this study to refer to the percentage of respondents who scored above the cut-off scores.

### 2.5. Statistical analysis

This is a descriptive study of the applicable rates of gambling disorder, showing the number and percentage of people who have participated in gambling in the 2 cities. A chi-square test was conducted for differences in the number of applicable cases for each of the gambling disorder scales for gender and region. In addition, the distribution of PACs for the 4 screening tests was illustrated in a special chart.

### 2.6. Ethical approval

Permission to conduct this study was obtained from the Nara Medical University Ethics Committee (Permission No. 2892. on 2021-3-26).

## 3. Results

### 3.1. Basic Attributes

Table 1 shows the number of persons by age group overall and by age group in each region. Numbers in parentheses indicate the percentage of males. Comparisons with the age and sex percentages in the total population of Osaka and Fukuoka prefectures at the time of the survey indicated that the respondents of this survey were represented as a sample that roughly matched the age and gender percentages of each region.

**Table 1.**
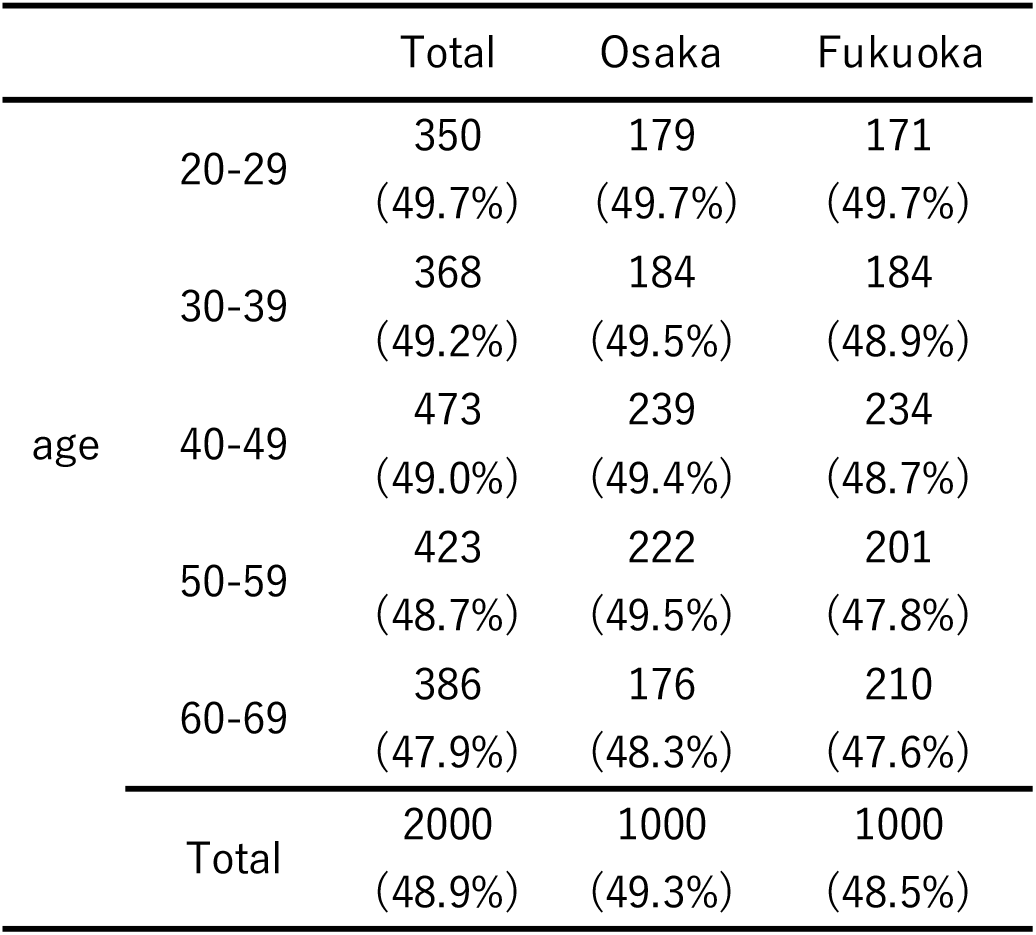
Basic attributes of participants

### 3.2. Profiles of gambling type

Table 2 shows the number and percentage of legal gambling participants in Japan. Each gamble is listed in descending order of total number of participants, with the inner number of gamblers shown as either experience within the past year or experience more than one year ago. The gambling activity with the largest number of participants was the lottery in which more than half of the respondents had participated. The second most popular gambling activity was PACHINKO followed by horse race as the third most popular gambling activity and sports promotion lottery as the fourth most popular. About 10% of the respondents had participated in other public races. About 10% of the respondents had participated in casino outside Japan, indicating that their experience was similar to that of those who had participated in boat race and bicycle race. By region, Osaka Prefecture had a higher number of participants in horse race than Fukuoka Prefecture, but the number of participants in other gambling activities was about the same.

**Table 2.** Number and percentage of people participating in gambling activity

|  |  | Total number of<br>experienced | Betting experience<br>within the past year | Betting experience<br>over a year ago |
| --- | --- | --- | --- | --- |
| lottery | total | 1169 (58.5%) | 580 (29.0%) | 589 (29.5%) |
|  | Osaka | 580 (58.0%) | 291 (29.1%) | 289 (28.9%) |
|  | Fukuoka | 589 (58.9%) | 289 (28.9%) | 300 (30.0%) |
| PACHINKO | total | 844 (42.2%) | 246 (12.3%) | 598 (29.9%) |
|  | Osaka | 438 (43.8%) | 133 (13.3%) | 305 (30.5%) |
|  | Fukuoka | 406 (40.6%) | 113 (11.3%) | 293 (29.3%) |
| horse race | total | 674 (33.7%) | 294 (14.7%) | 380 (19.0%) |
|  | Osaka | 379 (37.9%) | 173 (17.3%) | 206 (20.6%) |
|  | Fukuoka | 295 (29.5%) | 121 (12.1%) | 174 (17.4%) |
| sports promotion lottery | total | 436 (21.8%) | 234 (11.7%) | 202 (10.1%) |
|  | Osaka | 225 (22.5%) | 125 (12.5%) | 100 (10.0%) |
|  | Fukuoka | 211 (21.1%) | 109 (10.9%) | 102 (10.2%) |
| boat race | total | 299 (15.0%) | 110 ( 5.5%) | 189 ( 9.5%) |
|  | Osaka | 131 (13.1%) | 58 ( 5.8%) | 73 ( 7.3%) |
|  | Fukuoka | 168 (16.8%) | 52 ( 5.2%) | 116 (11.6%) |
| casino outside Japan | total | 244 (12.2%) | 43 ( 2.2%) | 201 (10.1%) |
|  | Osaka | 134 (13.4%) | 24 ( 2.4%) | 110 (11.0%) |
|  | Fukuoka | 110 (11.0%) | 19 ( 1.9%) | 91 ( 9.1%) |
| bicycle race | total | 215 (10.8%) | 96 ( 4.8%) | 119 ( 6.0%) |
|  | Osaka | 100 (10.0%) | 51 ( 5.1%) | 49 ( 4.9%) |
|  | Fukuoka | 115 (11.5%) | 45 ( 4.5%) | 70 ( 7.0%) |
| motorcycle race | total | 169 ( 8.5%) | 76 ( 3.8%) | 93 ( 4.7%) |
|  | Osaka | 66 ( 6.6%) | 37 ( 3.7%) | 29 ( 1.7%) |
|  | Fukuoka | 103 (10.3%) | 39 ( 3.9%) | 64 ( 6.4%) |

Regarding the number of participants in the past year, the lottery had the largest number of participants, followed by horse race PACHINKO and sports promotion lottery in second, third, and fourth, respectively. By region, the order of participation did not change, but the number of respondents who participated in horse races was higher in Osaka Prefecture than in Fukuoka Prefecture. Regarding gambling activities in which respondents reported participating at least once a week within the past year, the overall order of participation was lottery, horse race, PACHINKO and sports promotion lottery in order of the number of participants. By region, PACHINKO was second and the horse race third in Fukuoka Prefecture, showing regional characteristics.

### 3.3. Number and percentage above each questionnaireʼs cut-off scores

Table 3 shows the cumulative number of applicable cases and PACs for the four gambling disorder measures as a whole, by gender, and for each region. For the PACs by gender and region, the calculations were made for males (978), females (1,022), and Osaka and Fukuoka (1,000 each), respectively, as a whole. For each cumulative number of cases and PAC, SOGS calculations are based on lifetime responses to one’s gambling activity, while PGSI, LieBet, and DSM-5 calculations are based on responses to one’s gambling activity in the past 12 months. The cumulative number of patients with SOGS cut-off score (5 points) or higher was 205 (164 males and 41 females), with a PAC of 10.3% (16.3% males and 4.0% females). The cumulative number of patients with the cut-off score (1 point) or higher for the LieBet was 97 (78 males, 19 females), and the PAC was 4.9% (8.0% males, 1.9% females). 61, 14 females), with a PAC of 3.8% (6.2% male, 1.4% female). Looking at the overall cumulative number of applicable cases, SOGS had the highest number of cases, followed by PGSI, LieBet, and DSM-5, in that order, with the lowest number of cases. A Cochran’s Q test on these results revealed a significant difference in the cumulative number of applicable cases among the scales (*Q* = 156.38, *df*=3, *p* < .001). Multiple comparisons revealed significant differences between SOGS and each of the remaining scales. Differences were also found between the PGSI, LieBet, and DSM-5 in the number of applicable cases. Next, Cochran’s Q-tests were conducted to determine if there were any differences in the cumulative number of PGSI, LieBet, and DSM-5 cases calculated based on responses regarding one’s gambling activity over the past 12 months. The results revealed that there was a difference in the cumulative number of cases among the three scales (*Q* = 38.65, *df*=2, *p* < .001).

**Table 3.** Number and percentage above cut-off score (total, by gender, by region).

| | SOGS ( $\geq 5$ )<br>lifetime | PGSI ( $\geq 8$ )<br>12 months | LieBet ( $\geq 1$ )<br>12 months | DSM ( $\geq 4$ )<br>12 months |
| --- | --- | --- | --- | --- |
| all <sup>†</sup> | 205 (10.3%) | 134 ( 6.7%) | 97 ( 4.9%) | 75 ( 3.8%) |
| male | ** [ 164 (16.8%) | ** [ 105 (10.7%) | ** [ 78 ( 8.0%) | ** [ 61 ( 6.2%) |
| female | 41 ( 4.0%) | 29 ( 2.8%) | 19 ( 1.9%) | 14 ( 1.4%) |
| Osaka | [ 119 (11.9%) | 73 ( 7.3%) | 56 ( 5.6%) | [ 47 ( 4.7%) |
| Fukuoka | * [ 86 ( 8.6%) | 61 ( 6.1%) | 41 ( 4.1%) | * [ 28 ( 2.8%) |

Note. \*\* = $p < .001$ , \* = $p < .05$ , <sup>†</sup> : All combinations except between LieBet and DSM were significant. Values in parentheses shows percentage above cut off score (PAC).

Comparisons between the scales also revealed differences in the cumulative number of cases for the PGSI and the LieBet, and for the PGSI and the DSM-5. Next, a chi-square test was conducted to determine whether there were differences in the cumulative number of participants for each scale in men and women, and the results showed that the SOGS (*χ*^2^(1) = 88.41, *p* < .001, *ϕ* = -0.21), PGSI (*χ*^2^(1) = 49.88, *p* < .001, *ϕ* = - 0.16), LieBet (*χ*^2^(1) = 40.51, *p* < .001, *ϕ* = -0.14), and DSM-5 (*χ*^2^(1) = 32.80, *p* < .001, *ϕ* = -0.13), all four scales showed significant differences. Residual analysis indicated that the cumulative number of male subjects was significantly higher than the cumulative number of female subjects in all four scales.

A chi-square test was conducted to determine whether there was a difference in the cumulative number of participants for each scale between Osaka and Fukuoka, and significant differences were found between the SOGS (*χ*^2^(1) = 5.92, *p* = .02, *ϕ* = -0.54) and DSM-5 (*χ*^2^(1) = 5.00, *p* = .03, *ϕ* = -0.05). The residual analysis revealed significant differences between the two scales. Residuals analysis showed that the cumulative number of participants in Osaka was significantly higher than that in Fukuoka for the two scales, while no significant differences were found for the PGSI (*χ*^2^(1) = 1.15, *p* = .28, *ϕ* = -0.02) and LieBet (*χ*^2^(1) = 2.44, *p* = .12, *ϕ* = -0.04). No significant difference was found.

### 2.4. Comparison between scales

Figure 1 shows the PAC of four gambling disorder measures. The numbers in the circles represent scores, and the percentages near the circles represent PAC. The number of people who scored on each scale is generally proportional to the size of the circle, and the arc at the top indicates the population of those who scored zero on each scale. Standard cut-off scores for each scale are indicated by bold lines. We call this chart “dango chart” because it looks like the shape of “dango”, a traditional Japanese rice sweet.

**Figure 1.**
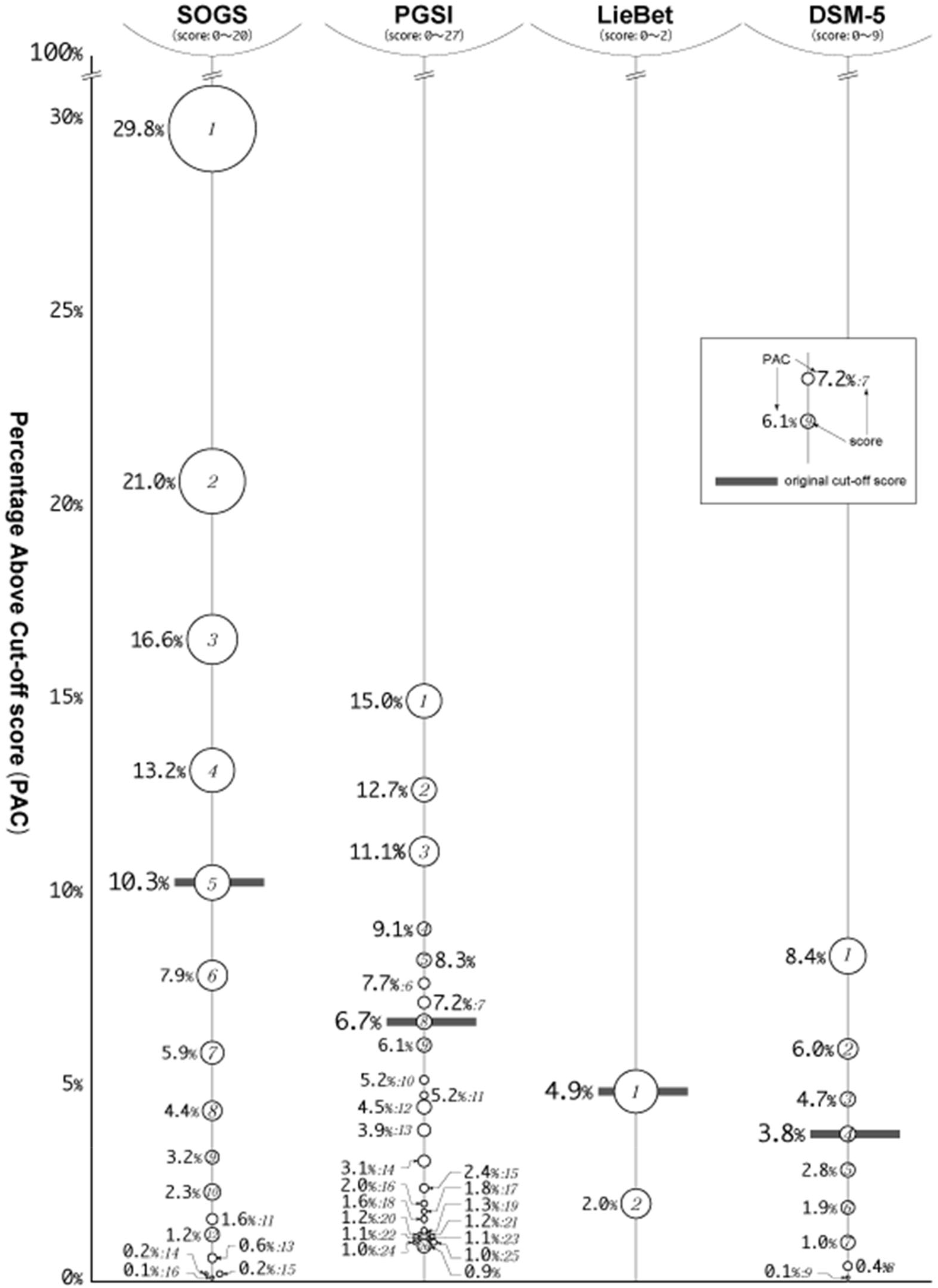
PAC distribution across the 4 gambling disorder measures (dango chart) The size of each circle is proportional to the number of people in that score.

Although it should be noted that SOGS is the lifetime rate and the other three are the rates for the past year, there is a nearly threefold difference in the PAC at the standard cut-off scores between DSM-5 (3.8%) and SOGS (10.3%), indicating that SOGS (lifetime) is more likely than SOGS (one year). Even excluding SOGS (lifetime), there is a nearly two-fold difference between DSM-5 and PGSI. In addition, while 70.2% (100-29.8) of the applicable respondents had a score of 0 for SOGS, there was a large difference of 95.1% (100-4.9) for LieBet.

## 4. Disucussion

### 4.1. Discussion of each result

#### 4.1.1. Profiles of gambling type

Regarding the experience of participating in legal gambling activities in Japan, the overall results showed that lottery had the largest number of participants, followed by PACHINKO, horse race and sports promotion lottery in that order. Only about 10% of all respondents had participated in other public races and casinos overseas.

PACHINKO is the most popular gambling activity in Japan, with the largest market size (Headquarters for Social Productivity, 2021). The results of the Chiba City (2018) survey and the 2020 national prevalence study (Matsushita, Nitta, & Toyama, 2021) were similar for each gambling activity, and we believe that we were able to show the actual situation regarding the experience of participating in gambling activities that are not illegal in Japan.

In terms of participation in gambling within the past year, horse race surpassed PACHINKO albeit only slightly. This result may be due to the self-restraint of PACHINKO parlors during the Corona Disaster and the increase in online horse betting opportunities. By region, the number of respondents who selected horse race and PACHINKO differed between Osaka and Fukuoka prefectures, possibly due to the number of racetracks and off-track ticket offices existing in those areas.

The number of respondents who have participated in casinos overseas is lower than the number of respondents who have participated in lottery, PACHINKO, horse race, and sports promotion lottery. It is necessary to continue to collect data on how the participation experience will change in the future as Japan attracts more casinos.

#### 4.1.2. Number and percentage above each questionnaireʼs cut-off scores

For the PAC of SOGS, the figure was about three times higher than the lifetime prevalence (3.6%) obtained in the 2017 national prevalence study (Higuchi and Matsushita, 2017). For the PAC of PGSI, the figure was slightly higher than the point in time prevalence (4.8%) obtained in the Chiba City (2018) survey and about four times higher than the point in time prevalence (1.6%) obtained in the 2020 national prevalence study (Matsushita, Nitta, & Toyama, 2021). The reason for the very high figures compared to the national prevalence study may be due to the fact that the survey was conducted in Osaka and Fukuoka prefectures, which have large populations, as well as the use of an online survey. However, it is also possible that the results of the national prevalence study were influenced by the use of interviews and a portion of mail surveys, resulting in low estimates of lifetime prevalence and point-prevalence. We plan to continue conducting the online survey in the future, and we believe it is necessary to take these points into account when reviewing the survey results.

Comparison of the cumulative number of applicable cases and PAC among scales revealed that SOGS was the highest, followed by PGSI, LieBet, and DSM-5, in that order. The PGSI was also found to have the highest PAC when compared to the PGSI, LieBet, and DSM-5, which asked respondents to report their gambling activity in the past 12 months. SOGS was found to screen for more possible gambling activity compared to the other measures.

#### 4.1.3. Comparison between scales (dango chart)

In the comparison between scales (Figure 1), the characteristics of each scale are illustrated by differences in the distribution of PAC. The number of respondents who scored 1 or more points on each scale (PAC at 1 point) was high for the SOGS at 29.8% and for the PGSI at 15.0%, while it was low for the DSM-5 at 8.4% and for the LieBet at 4.9%. In other words, the SOGS and PGSI had more positive responses to any of the items in the scale, while the DSM-5 and LieBet had more positive responses to none of the items (more people scored zero). The SOGS and PGSI can detect mild to moderate conditions depending on the cut-off scores. And the dango chart allows us to visually grasp the relationship between the scales in the same population.

By administering multiple scales to the same study subjects and showing the distribution of PACs for each scale as shown in Figure 1, it was possible to set appropriate cut-off scores and examine the possibility of obtaining the “severity” that each scale possesses.

### 4.2. General Discussion

This study presents the number and percentage of people in Japan who have participated in legal gambling, the applicable rates for the 4 gambling disorder screening tests by region and gender, and the distribution of PACs among the several screening tests. This is the first study to compare gambling disorder rates between a region where an IR is planned to open and a region where it is not, and to administer the 4 screening tests to the same subjects at the same time.

Although the survey did not find any remarkable regional differences in the number and percentage of participants, it is an important clinical and policy discussion point whether regional differences will emerge with the opening of casinos in Japan in the future. The finding of significant gender differences in the number of participants in the screening test was consistent with previous studies.

In the results comparing PACs in the PGSI, LieBet, and DSM-5, all PAC points are concentrated in a narrow range in the LieBet and DSM-5. This reveals that the LieBet and DSM-5 are measures that screen for more severe groups.

This study has several limitations. First, this study is an online survey. Previous studies have suggested that online surveys tend to estimate applicable rates higher than other survey methods (Nower, Volberg & Caler, 2017). Second, SOGS in this study calculates the cumulative lifetime number of applicable persons. It should be noted that this may result in a higher applicable person rate than the other 3 tests. This study shows the applicable gambling disorder rates in a region where an IR is planned to be opened and in a region where not, and is expected to contribute as a baseline study for measuring the impact of the opening of an IR. In addition, this study administered several gambling disorder screening tests to the same subjects in the same region and illustrated them in a chart format named the dango chart.

This visualized the changes in the applicable person rate when the cutoff values of the screening tests were changed (within-test comparisons) and the differences in the distribution of the applicable person rate across tests (between-test comparisons). We think that the dango chart is a useful presentation method for comparing several different tests.

Although the online survey may cause bias in the direction of overestimation, by continuing to use the same measurement method, our research project will be able to reach its ultimate goal of observing changes in the applicable rate before and after the opening of the IR.

## Data Availability

All data from this study are available upon reasonable request to the authors.

## Notes

**Role of funding source** This work was supported by JSPS KAKENHI Grant Number JP17K09092 and JP20K14237.

**Conflict of interest** The authors have no conflicts of interest to this manuscript.

### Competing Interest Statement

The authors have declared no competing interest.

### Funding Statement

This work was supported by JSPS KAKENHI Grant Number JP17K09092 and JP20K14237.

